# Seasonally Variant Gene Expression in Full-Term Human Placenta

**DOI:** 10.1101/2020.01.27.20018671

**Authors:** Danielle A. Clarkson-Townsend, Elizabeth Kennedy, Todd M. Everson, Maya A. Deyssenroth, Amber A. Burt, Ke Hao, Jia Chen, Machelle Pardue, Carmen J. Marsit

## Abstract

Seasonal exposures influence human health and development. The placenta, as a mediator of the maternal and fetal systems and a regulator of development, is an ideal tissue to understand the biological pathways underlying relationships between season of birth and later life health outcomes. Here, we conducted a transcriptome-wide association study of season of birth in full-term human placental tissue to evaluate whether the placenta may be influenced by seasonal cues. Of the analyzed transcripts, 583 displayed differential expression between summer and winter births (FDR q<0.05); among these, *BHLHE40, MIR210HG*, and *HILPDA* had increased expression among winter births (Bonferroni p<0.05). Enrichment analyses of the seasonally variant genes between summer and winter births indicated over-representation of transcription factors HIF1A, VDR, and CLOCK, among others, and of GO term pathways related to ribosomal activity and infection. Additionally, a cosinor analysis found rhythmic expression for approximately 11.9% of all 17,664 analyzed placental transcripts. These results suggest that the placenta responds to seasonal cues and add to the growing body of evidence that the placenta acts as a peripheral clock, which may provide a molecular explanation for the extensive associations between season of birth and health outcomes.

## INTRODUCTION

Seasonal changes in weather(1), pathogens(2), pollutants(3), nutrient contents(4), and photoperiod(5) can influence human health. Associations between season of birth and health outcomes such as autoimmune disease(6), myopia(7), and even lifespan(8), are well-documented(9), and developmental photoperiod is one of the proposed mechanisms behind these relationships(10). However, annual fluctuations in conception and birthrates can confound associations between season of birth and health outcomes(11, 12). Furthermore, maternal sociodemographic variables, such as maternal education, marital status, and socioeconomic status, are also associated with season of birth(11, 13), which may drive some of the seasonal disparities in health outcomes. However, the associations between season of birth and health outcomes may not solely be the result of possible bias from maternal characteristics; a large study comparing multiple births within mothers (to control for maternal sociodemographic characteristics), found that birth outcomes still displayed a seasonal pattern(11), suggesting an underlying biological effect of environmental factors that vary with season. While photoperiod is almost certainly not the only driver of seasonal differences, the influence of environmental photoperiod on the biological clock may be a key factor in the relationship between seasonality and disease.

Light is a salient *zeitgeber* (“time giver”) of the circadian clock, and annual changes (in non-equatorial regions) in photoperiod act as a reliable seasonal cue for the clock(14, 15). When light reaches the eye, the light signal is relayed to the suprachiasmatic nucleus of the brain, the central clock, which then entrains other tissues, or peripheral clocks, to the external environment. With the widespread use of indoor lighting, the majority of people in the U.S. are not reliant on outdoor light exposure for illumination; however, the amount of UV light exposure and blood levels of the hormone Vitamin D fluctuate with photoperiod(16), with a peak in the summer and trough in the winter. Melatonin production also fluctuates seasonally(17), roughly opposite of Vitamin D with a peak in the winter and trough in the summer. In addition to rhythms in hormone production, rhythmic patterns of gene expression may also underlie season and health relationships(18). Studies in humans and other animals have found seasonal patterns of gene expression in tissues such as blood(18-20) and adipose tissue(18, 21), although some of these differences may also reflect seasonal changes in cell populations(22). These rhythms can vary by tissue type and share pathways with the core circadian clock system(23).

The placenta is an important intermediary between maternal photoperiod and fetal development; as a tissue with neuroendocrine properties, the placenta transports and synthesizes hormones related to light exposure, such as melatonin and the active form of vitamin D(24-26). Given these characteristics, we hypothesize that the placenta responds to seasonal cues such as photoperiod. To test this hypothesis, we investigated transcriptome-wide placental gene expression from full term (≥37 weeks) pregnancies delivered during different seasonal photoperiods.

## MATERIALS AND METHODS

### Placental sequencing

The analysis utilized placental sequencing data from the Rhode Island Child Health Study (RICHS) cohort, as previously described(27-29). RNA-seq was performed on a subset of placenta samples (n=200) from the RICHS cohort with the Illumina HiSeq 2500 platform to measure gene expression, as previously described(29). This data is available in dbGaP (phs001586.v1.p1). Briefly, FASTQ files were checked for quality and all reads had a phred score > 30. The reads were trimmed using “seqtk”(30) with an error threshold of 0.01 and no minimum length filter. These trimmed FASTQ files were aligned to the human reference genome (Homo sapiens GRCh37.75.gtf version hg19) using STAR(31) v2.4.0g1 with a 15bp minimum overhang and minimum mapped length for chimeric junctions. Counts were mapped from the BAM file using “subread”(32) v1.5.0-p1. Of this cohort, 1 sample was removed due to withdrawn consent and 2 samples removed due to mismatched sex, leaving a total of 197 placental samples included in this analysis.

### Exposure characterization

Demographic and medical information were provided via structured interviews and standardized medical record abstraction by trained staff. Month of birth was used to characterize the season of birth into one of 4 photoperiod categories: SS (summer solstice), FE (fall equinox), WS (winter solstice), and SE (spring equinox); these categories were based on the midpoints between solstice and equinox dates and rounded to the nearest month to bin by light exposure, where SS included those born from the months of May through July, FE included those born from August through October, WS included those born from November through January, and SE included those born from February through April (**Figure 1**), similar to other studies(33, 34). Placenta samples were collected from 2009-2013 during the daytime hospital hours of 8AM to 4PM. Possible confounders were evaluated, including: method of delivery (C-section vs. vaginal), maternal adversity, maternal pre-pregnancy smoking, infant gestational age, infant year of birth, sex of the infant, infant birthweight category, infant birthweight z-score, maternal BMI, maternal weight gain during pregnancy, maternal gestational diabetes, maternal night shift work, maternal age, maternal race, and hour of sample collection. One sample with missing data for the maternal pre-pregnancy smoking variable was recoded to “No”. The maternal adversity cumulative variable(27) was created to control for maternal sociodemographic characteristics based on maternal education (high school or less = +1), maternal insurance (public, self-pay/ other, or none = +1), maternal support (single and no support = +1), household income adjusted for household size and based on the federal poverty level during the year of birth (below poverty level = +1), and household size (>6 people in same household = +1); this cumulative variable was split into 3 levels (+0, +1, or ≥+2), with higher score indicating greater adversity.

**Figure 1.**
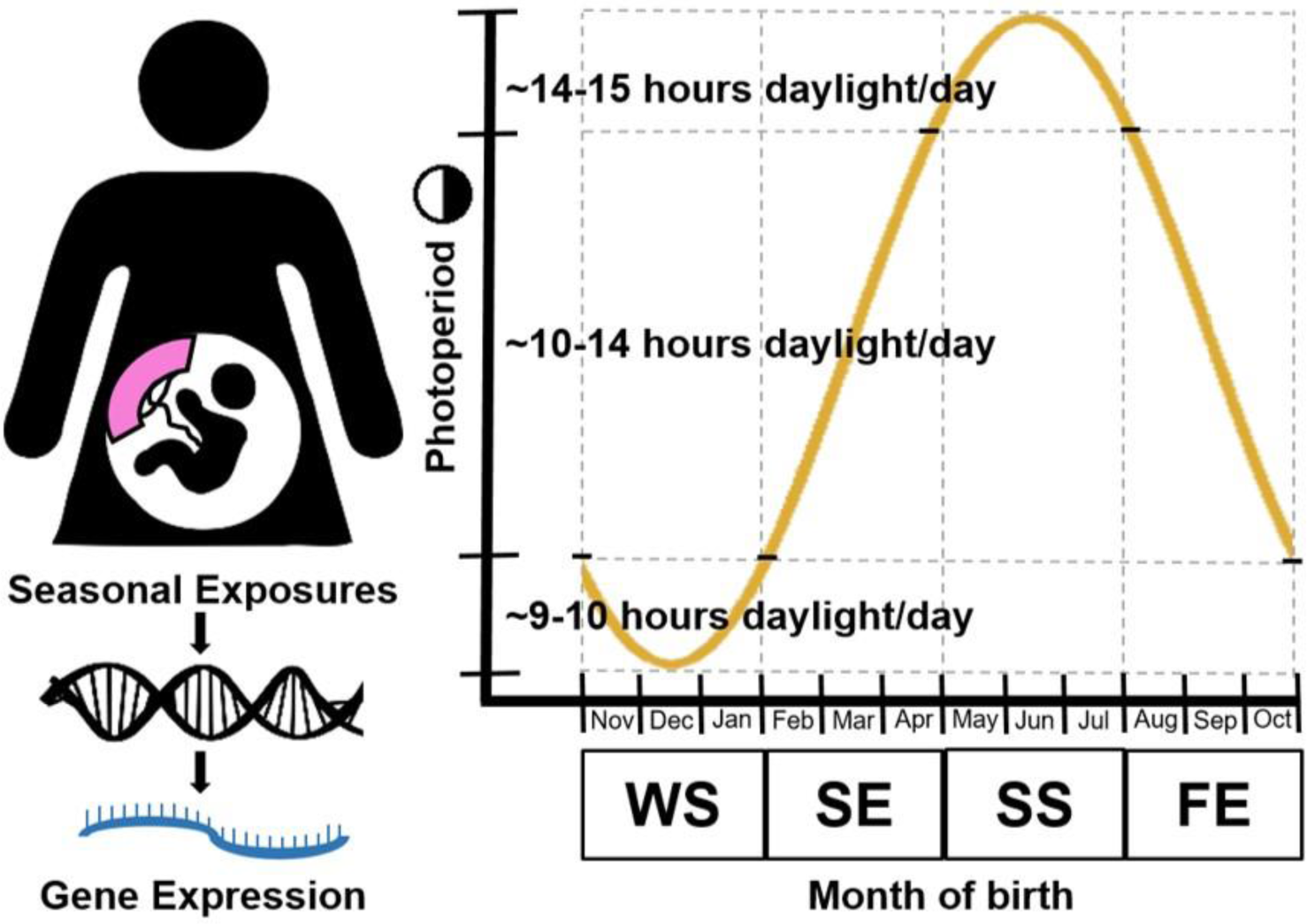
Illustration of overall study concept and categorization of seasonal exposure groups based on approximate annual photoperiod fluctuations around Providence, Rhode Island, USA (41.81° N, 71.41° W), where the RICHS study enrollment took place.

### Differential expression analysis

RNA-seq count data was read into R and differential expression (DE) analysis was performed using the *DESeq2* package(35). Before analysis, low count data were filtered from the dataset using a threshold to exclude transcripts with <2 counts for 10% of samples. To control for unwanted heterogeneity due to other batch effects or unmodeled factors and to improve the reproducibility of the results, we also employed surrogate variable analysis(36) using the *sva* package(37) in R. To estimate surrogate variables, counts were normalized and the surrogate variables were estimated with the Buja and Eyuboglu (“be”) permutation method(38) using 200 iterations and then iteratively reweighted in svaseq(39). Normalized gene expression with seasonal photoperiod as the exposure was modeled using *DESeq2*, adjusting for maternal adversity and precision variables that included sex of the infant, method of delivery, gestational age, maternal pre-pregnancy smoking, infant ‘s year of birth, sequencing batch, and the first two estimated surrogate variables. Overall effect of season (across all photoperiod categories) was first evaluated using the likelihood ratio test (LRT) method. Because the primary seasons of interest were those with the longest and shortest photoperiods, the Wald test method was used to compare WS and SS (reference group). However, for visualization of how expression changed across seasons, FE and SE were also compared to SS in subsequent sensitivity analyses. To control for false discovery rate (FDR) due to multiple comparisons, we employed the Benjamini and Hochberg (BH) adjustment method(40) and the more conservative Bonferroni adjustment method.

### Cosinor rhythmicity analyses

To evaluate whether placental gene expression displayed rhythmicity, we conducted a cosinor analysis of all placental transcripts that met the count threshold using the *cosinor* package in R(41, 42). A cosinor model was fit to the variance standardized transformation (VST) of gene expression for each transcript using methods laid out by Tong (1976)(41). Briefly, the cosinor function was applied to each transcript with the general formula:

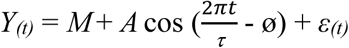

Where the outcome *Y(t)* is the variance-standardized (VST) gene expression for a gene transcript at time *t* (birth month), *M* is the intercept of the fitted curve, A is the amplitude of the curve, *τ* is the fixed period length (12 months), ø is the acrophase of the curve, and ε is an error term with mean 0. In this way, models with a time period (*τ*) of 12 were fitted to each transcript to test whether they displayed a significant amplitude and/or acrophase compared to baseline using the consinor.lm function of the *cosinor(42)* package with the general code:

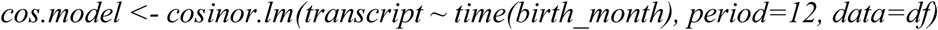

To test the fit of the cosinor model compared to the intercept, global F tests were performed and results controlled for multiple comparisons using the BH or Bonferroni method; amplitude or acrophase were considered significant if FDR q<0.05.

### Bioinformatic analyses

To identify transcription factors and pathways over-represented amongst the SS and WS photoperiod, representing the longest and shortest photoperiods, DE genes (FDR q<0.05) were analyzed for functional enrichment against references gene lists within the ChEA 2016, KEGG 2019, and GO 2018 databases using enrichR(43). Enrichment significance was computed using Fisher ‘s exact test and considered significant if resulting FDR q<0.05.

## RESULTS

Of the 197 placental samples included in the analysis, 60 (30%) were delivered during the SS season, 57 (29%) during the FE season, 31 (16%) during the WS season, and 49 (25%) during the SE season (**Table 1, Figure 1**). Infant birthweight group, infant birthweight z-score, maternal BMI, maternal weight gain during pregnancy, maternal gestational diabetes, maternal night shift work, maternal age, maternal race, and hour of sample collection did not significantly differ by seasonal category (**Table 1, Supplemental Figure 1**). Maternal adversity significantly differed across seasonal categories (p=0.01) and was included in the final model. The final DE model adjusted for maternal adversity, sex of the infant, method of delivery, gestational age, maternal pre-pregnancy smoking, infant ‘s year of birth, sequencing batch, and the first two estimated surrogate variables, with season of birth as the exposure of interest and normalized gene expression the outcome of interest (**Table 1**).

**Table 1.** Demographic characteristics evaluated for inclusion in the final model testing differential expression. To evaluate statistical difference of variables across birth categories, 1-way ANOVA was used for continuous variables and chi-square or Fisher ‘s exact tests were used for categorical variables.

| Variable | Season of birth |  |  |  | p-val |
| --- | --- | --- | --- | --- | --- |
|  | SS<br>(n=60) | FE<br>(n=57) | WS<br>(n=31) | SE<br>(n=49) |  |
| **C-section, n ( %)** | 21 (35.0) | 11 (19.3) | 9 (29.0) | 16 (32.7) | 0.264 |
| **Maternal adversity, n (%)** |  |  |  |  | 0.013 |
| 0 | 30 (50.0) | 42 (73.7) | 21 (67.7) | 38 (77.6) |  |
| 1 | 15 (25.0) | 5 ( 8.8) | 7 (22.6) | 8 (16.3) |  |
| ≥2 | 15 (25.0) | 10 (17.5) | 3 (9.7) | 3 (6.1) |  |
| **Maternal pre-pregnancy smoking, n (%)** | 6 (10.0) | 3 ( 5.3) | 2 ( 6.5) | 5 (10.2) | 0.757 |
| **Gestational age** |  |  |  |  | 0.190 |
| 37-38 weeks | 15 (25.0) | 8 (14.0) | 7 (22.6) | 10 (20.4) |  |
| 39 weeks | 34 (56.7) | 33 (57.9) | 18 (58.1) | 20 (40.8) |  |
| 40-41 weeks | 11 (18.3) | 16 (28.1) | 6 (19.4) | 19 (38.8) |  |
| **Year of birth** |  |  |  |  | 0.055 |
| 2009-2010 | 34 (56.7) | 27 (47.4) | 12 (38.7) | 13 (26.5) |  |
| 2011 | 13 (21.7) | 16 (28.1) | 9 (29.0) | 14 (28.6) |  |
| 2012-2013 | 13 (21.7) | 14 (24.6) | 10 (32.3) | 22 (44.9) |  |
| **Male infant, n (%)** | 27 (45.0) | 30 (52.6) | 14 (45.2) | 25 (51.0) | 0.815 |
| **Birthweight Group, n (%)** |  |  |  |  | 0.660 |
| Small for gestational age | 8 (13.3) | 11 (19.3) | 7 (22.6) | 6 (12.2) |  |
| Average for gestational age | 33 (55.0) | 30 (52.6) | 19 (61.3) | 29 (59.2) |  |
| Large for gestational age | 19 (31.7) | 14 (28.6) | 5 (16.1) | 14 (28.6) |  |
| **Birthweight z-score, mean (SD)** | 0.24 (1.18) | 0.32 (1.34) | -0.20 (1.16) | 0.23 (1.24) | 0.281 |
| **Maternal BMI, mean (SD)** | 26.67 (6.35) | 27.58 (7.44) | 24.14 (4.97) | 25.89 (5.85) | 0.104 |
| **Maternal Weight Gain (kg), mean (SD)** | 15.32 (5.58) | 14.11 (6.08) | 13.86 (6.61) | 14.91 (6.30) | 0.622 |
| **Maternal Gestational Diabetes (yes), n (%)** | 5 (8.3) | 6 (10.5) | 4 (14.3) | 4 (8.3) | 0.818 |
| **Maternal Night Shift Work (yes), n (%)** | 11 (21.2) | 11 (20.8) | 6 (20.7) | 11 (23.9) | 0.980 |
| **Maternal Age (yrs)** |  |  |  |  | 0.398 |
| 18-31 | 34 (56.7) | 24 (42.1) | 17 (54.8) | 23 (46.9) |  |
| 32-40 | 26 (43.3) | 33 (57.9) | 14 (45.2) | 26 (53.1) |  |
| **Maternal Race (white), n (%)** | 36 (64.3) | 45 (81.8) | 21 (70.0) | 38 (77.6) | 0.171 |
| **Hour Sample Collection, mean (SD)** | 11.51 (2.30) | 12.35 (2.37) | 12.45 (2.23) | 11.51 (2.47) | 0.083 |

After applying a threshold to filter out genes with counts <2 for 10% of samples, the number of transcripts modeled in the analysis decreased from the original set of 50,810 transcripts to a filtered set of 17,664 transcripts. In the resulting overall likelihood ratio test analysis of DE across all seasonal categories, 819 transcripts were considered significant after adjusting for multiple comparisons (FDR q<0.05, **Supplemental Table 1**), suggesting an overall effect of season. To measure expression differences between the specific seasonal categories of interest, the Wald test comparing the SS and WS groups found DE of 583 transcripts (FDR q<0.05), of which 574 were annotated to genes (**Figure 2, Supplemental Table 2**). Compared to SS, 347 transcripts had increased expression and 236 had decreased expression in WS. While not the primary aim of the study, DE results of the FE and SE seasonal groups compared to SS are also provided (**Supplemental Table 2, Supplemental Figure 2**).

**Table 2.** List of Bonferroni-significant (p<0.05) genes differentially expressed in SS vs WS placenta that also displayed rhythmicity in the cosinor analysis (FDR q<0.05).

| **Ensemble ID** | **Gene ID** | **Gene description** |
| --- | --- | --- |
| ENSG00000143222 | UFC1 | ubiquitin-fold modifier conjugating enzyme 1 [Source:HGNC Symbol;Acc:HGNC:26941] |
| ENSG00000134107 | BHLHE40 | basic helix-loop-helix family member e40 [Source:HGNC Symbol;Acc:HGNC:1046] |
| ENSG00000135245 | HILPDA | hypoxia inducible lipid droplet associated [Source:HGNC Symbol;Acc:HGNC:28859] |
| ENSG00000160072 | ATAD3B | ATPase family AAA domain containing 3B [Source:HGNC Symbol;Acc:HGNC:24007] |
| ENSG00000135441 | BLOC1S1 | biogenesis of lysosomal organelles complex 1 subunit 1 [Source:HGNC Symbol;Acc:HGNC:4200] |
| ENSG00000169860 | P2RY1 | purinergic receptor P2Y1 [Source:HGNC Symbol;Acc:HGNC:8539] |
| ENSG00000043591 | ADRB1 | adrenoceptor beta 1 [Source:HGNC Symbol;Acc:HGNC:285] |
| ENSG00000166167 | BTRC | beta-transducin repeat containing E3 ubiquitin protein ligase [Source:HGNC Symbol;Acc:HGNC:1144] |
| ENSG00000165775 | FUNDC2 | FUN14 domain containing 2 [Source:HGNC Symbol;Acc:HGNC:24925] |
| ENSG00000162408 | NOL9 | nucleolar protein 9 [Source:HGNC Symbol;Acc:HGNC:26265] |
| ENSG00000247095 | MIR210HG | MIR210 host gene [Source:HGNC Symbol;Acc:HGNC:39524] |
| ENSG00000186184 | POLR1D | RNA polymerase I and III subunit D [Source:HGNC Symbol;Acc:HGNC:20422] |
| ENSG00000100906 | NFKBIA | NFKB inhibitor alpha [Source:HGNC Symbol;Acc:HGNC:7797] |
| ENSG00000115211 | EIF2B4 | eukaryotic translation initiation factor 2B subunit delta [Source:HGNC Symbol;Acc:HGNC:3260] |
| ENSG00000163251 | FZD5 | frizzled class receptor 5 [Source:HGNC Symbol;Acc:HGNC:4043] |

**Figure 2.**
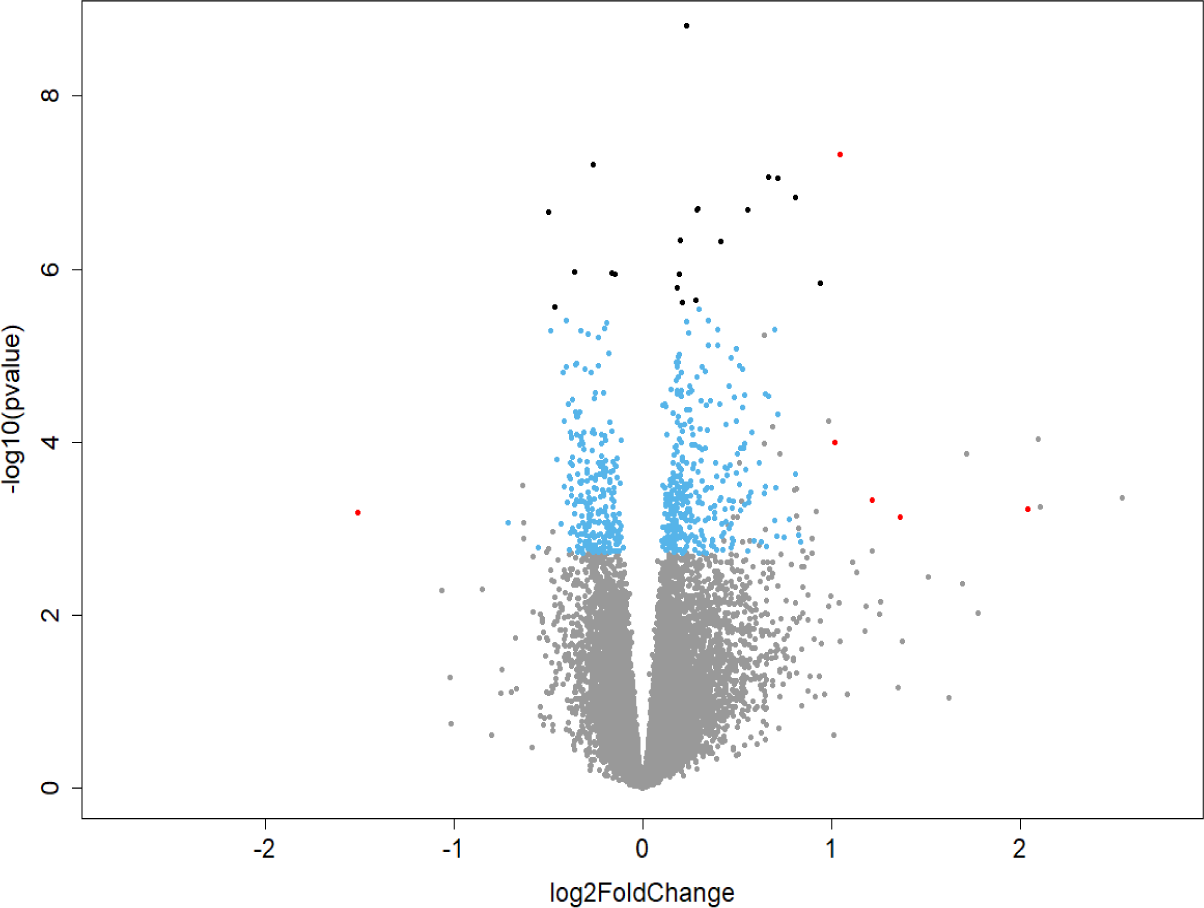
Volcano plot of results from the transcriptome wide association study of gene expression by birth season showing Wald test results comparing winter (WS) to the summer (SS) referent group. Transcripts are plotted by effect size estimate [Log2(Fold Change), x-axis] and statistical significance [-Log10(Pvalue), y-axis], with grey dots indicating non-significant FDR q≥0.05, blue dots indicating FDR q<0.05, black dots indicating Bonferroni p<0.05, and red dots indicating absolute log2(fold change)>1 and FDR q<0.05.

The cosinor analysis to measure gene expression rhythmicity by birth month found that of the 17,664 transcripts, 2,109 (11.9%) had significant seasonal amplitude and/or acrophase (FDR q<0.05) (**Figure 3, Supplemental Figure 3, Supplemental Table 3**). When the more conservative Bonferroni adjustment (p<0.05) was applied to the DE results, 15 genes were found to be significant in both the DE and cosinor analysis: *UFC1, BHLHE40, HILPDA, ATAD3B, BLOC1S1, P2RY1, ADRB1, BTRC, FUNDC2, NOL9, MIR210HG, POLR1D, NFKBIA, EIF2B4*, and *FZD5* (**Table 2**).

**Figure 3.**
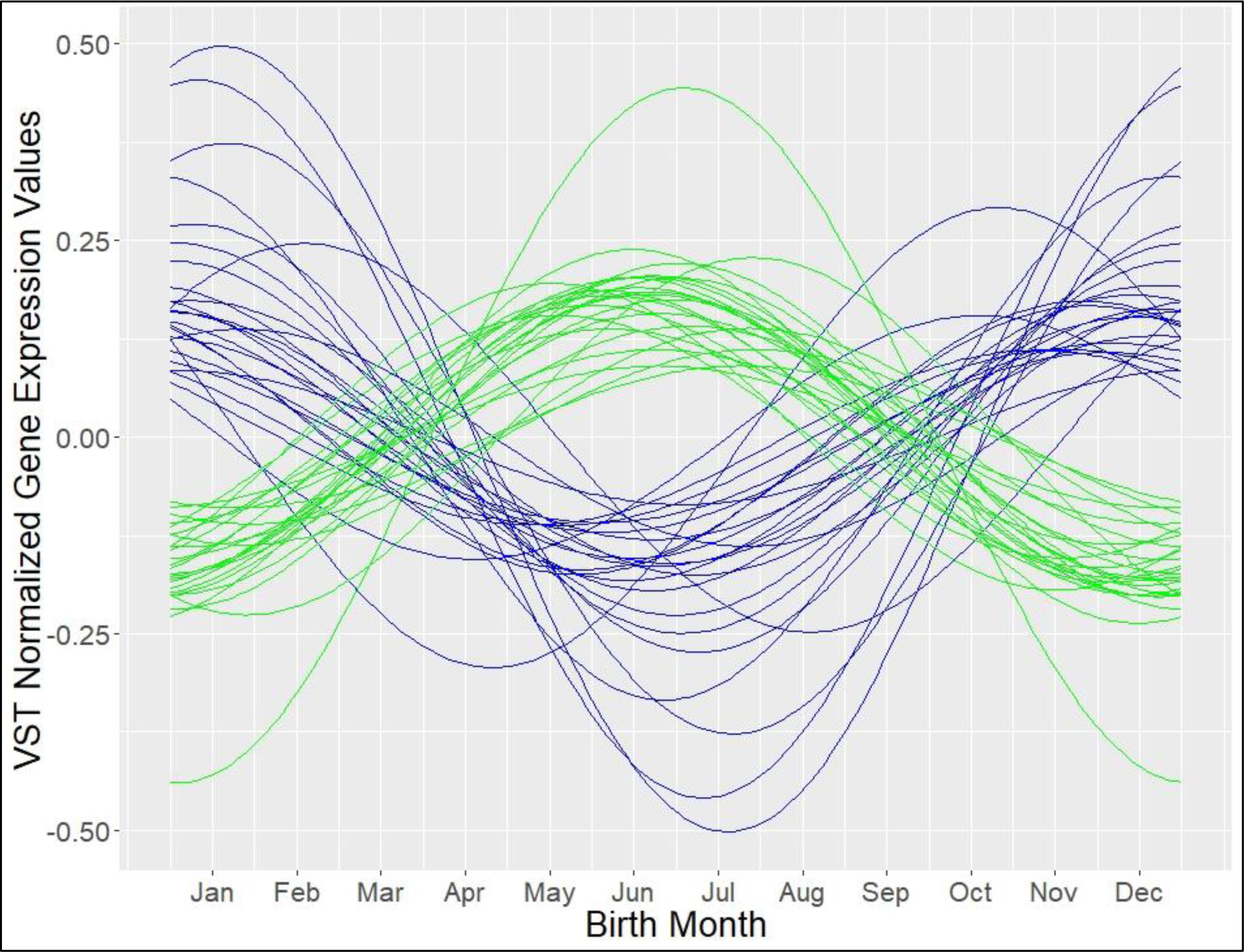
Graph showing the fitted curves of the top 25 upregulated and top 25 downregulated genes (FDR q<0.05) in the differential expression analysis comparing winter (WS) births to summer (SS) births when analyzed in the cosinor analysis. The green curves represent transcripts with peak expression in summer and the blue curves represent transcripts with peak expression in winter.

Examining the over-representation of transcription factor binding sites amongst the genes demonstrating DE between SS and WS using the ChEA database highlighted the transcription factors HIF1A, VDR, ELK1, TAL1, ELK3, FOXP1, CLOCK, NUCKS1, DMRT1, SPI1, DCP1A, ZNF217, and XRN2 to be over-represented (FDR q<0.05, **Supplemental Table 4**). These DE genes were also over-represented by the KEGG ribosome pathway and GO molecular and biological terms related to ribosomes, RNA processing, viral infection, and protein signaling to the endoplasmic reticulum (FDR q<0.05, **Supplemental Table 4**).

## DISCUSSION

The placenta, a transient organ that develops during pregnancy, plays a vital role in fetal health and development. Positioned at the boundaries of the developed maternal circadian system and the developing fetal circadian system, the placenta likely acts as a peripheral oscillator of the physiological circadian clock system, relaying maternal photic and other cues to the fetus. Our results found placental gene expression associated with seasons of birth, suggesting that the placenta may respond to seasonal cues such as photoperiod. While more studies are necessary, these findings provide support that seasonality of birth has underlying biological characteristics in humans that are separate from maternal demographic characteristics. This study adds to the growing body of evidence that the placenta acts as a peripheral clock(44).

Core clock genes such as *PER1, PER2, BMAL1, CLOCK, CRY1*, and *CRY2* are expressed in the placenta, but only *CLOCK* and *BMAL1* have so far been found to show rhythmic expression in full-term human placenta(45). Our results raise the possibility that the clock gene *BHLHE40* (also known as *DEC1*), which encodes a basic helix-loop-helix transcription factor that can bind to E-boxes, also exhibits rhythmicity in the placenta, as it was one of the top hits in the DE and cosinor analyses. Because BHLHE40 belongs to this family of transcription factors, it influences other circadian-related transcription factors such as BMAL1 through protein-protein binding or by competing for E-box binding sites(46, 47). BMAL1 oscillation in the SCN may encode seasonal photoperiod(48), and (in the northern hemisphere) a study found seasonal *BMAL1* expression displaying a trough around January and a peak around July(18); in line with these findings, our analysis found the expression of *BHLHE40* expression peaked around December-January (**Figure 3, Supplemental Figure 2**). Additionally, a SNP in BHLHE40 was found to be associated with placental abruption in full-term human placenta(49) and an *in vitro* experiment using a trophoblast cell line found rhythmic *BHLHE40* expression that was enhanced during hypoxic stress(50). These results support our findings of *BHLHE40* rhythmicity in the placenta and a possible role for this circadian clock gene in the placental hypoxia response.

In addition to *BHLHE40*, genes such as *UFC1, HILPDA, P2RY1, ADRB1, BTRC, MIR210HG, POLR1D, NFKBIA*, and *FZD5* appear to be rhythmic in the placenta (p<0.05) (**Table 2**). The representation of *BTRC* and *NFKBIA* is possibly due to seasonal inflammation, infection, and/or hypoxia and the NF-κB pathway(51). BTRC is an F-box protein that ubiquitinates NFKBIA for degradation, which disinhibits the NF-κB complex. In accordance, our results show inverse patterns of BTRC and NFKBIA expression, with BTRC peaking in the summer months and NKBIA peaking in the winter months. The representation of *MIR210HG* and *HILPDA* peaking in the winter months could also be due to seasonal hypoxia; HILPDA is involved in the hypoxia response(52) and *MIR210HG* is a long non-coding RNA precursor to miRNA-210 that is also activated during hypoxia(53). POLR1D, an RNA polymerase 1 and RNA polymerase 3 subunit that affects ribosomal synthesis and transcription(54), also had peak expression during the winter months. P2RY1 and ADRB1 both had peak expression during the summer months and function as G-protein coupled receptors; ADRB1 may play an important role in sleep behavior, with a mutation in ADRB1 found to decrease self-reported sleep need(55).

The results from the cosinor analysis of gene expression by birth month supported the findings of the seasonal DE analysis. When results (FDR q<0.05) from the SS vs WS DE analysis and the cosinor analysis were compared, 316 transcripts were represented in both analyses (**Supplemental Table 5**). The analysis of the overall 17,664 placental transcripts found 11.9% of them show seasonal rhythms, suggesting widespread seasonal rhythmicity in placental gene expression (**Supplemental Table 3**). Other analyses of seasonal and circadian gene expression have reported approximately 23%-43% of transcripts to be rhythmic(18, 56, 57), with transcript rhythmicity differing by tissue. There are no other studies of transcriptome-wide human placental rhythmicity, so more research is necessary to confirm genes with seasonal variation in the placenta.

Enrichment analyses indicated that the transcription factors HIF1A, VDR, ELK1, TAL1, ELK3, FOXP1, CLOCK, NUCKS1, DMRT1, SPI1, DCP1A, ZNF217, and XRN2 were over-represented amongst those genes that are DE between summer (SS) and winter (WS) births (**Supplemental Table 4**). Among these, HIF1A, TAL1, and CLOCK are members of the basic helix-loop-helix family of transcription factors and are associated with circadian rhythm regulation. HIF1A can also heterodimerize with BMAL1 and disruption of BMAL1 can induce HIF1A activity(58). During the early stages of placental development when conditions are hypoxic, HIF1A levels increase to promote trophoblast invasion; during later stages of pregnancy (>10-18 weeks), HIF1A levels decrease and stabilize(59); however, our analysis of full-term (≥37 weeks) placenta found over-representation of HIF1A binding sites associated with photoperiod (SS vs WS), suggesting environmental exposures may influence seasonal patterns in placental hypoxic signaling.

We also identified over-representation of binding sites for VDR, the vitamin D-receptor, lending credence to our results. As outdoor UV light exposure is highest during the summer months (SS) and lowest during the winter months (WS)(60), seasonal differences in genes controlled by this transcription factor are expected. Seasonal decreases in maternal UV light exposure could lead to decreased placental synthesis and transport of active vitamin D, altering VDR activity. VDR also heterodimerizes with RXR, another interactive component of the circadian clock, to bind vitamin D response elements in the genome and induce widespread gene expression.

The KEGG term and many of the GO terms associated with season of birth were related to ribosomes. Because ribosomes are responsible for translating mRNA into proteins, ribosomal abundance can affect rates of cell growth or transcriptional activity. The increased expression of ribosomal genes in winter births suggest that there may be seasonal differences in transcription rates. The GO term results of infection-related pathways possibly reflect the increased exposure to viral pathogens and infections during the winter months or a seasonally programmed ability to respond to infection.

There are some limitations to the study and results should be interpreted cautiously. Because temperature and photoperiod are so tightly linked, some of our results may be due to temperature (or other seasonally fluctuating exposures) rather than being exclusively driven by photoperiod. The changes in gene expression are possibly due, at least in part, to seasonal changes in underlying placental cell populations. Additionally, while we utilized surrogate variable analysis to control for unknown heterogeneity in samples, there may be false positives and residual confounding from unmodeled factors. While conception rates fluctuate seasonally, this effect may not have a large influence on the results as all samples were from full-term (≥37weeks), singleton pregnancies free from congenital abnormalities, infant birthweight z-scores did not significantly differ by season, and the model adjusted for maternal sociodemographic characteristics. Strengths of this study include the use of full-term human placenta, the use of a conservative count threshold for filtering transcripts used in the analyses, and the use of cosinor analyses to further confirm and characterize transcript rhythmicity by birth month. More research is necessary to elucidate the role of the placenta in circadian signal arbitration between the maternal and developing fetal systems.

In conclusion, we report seasonal variation in gene expression of human placental tissue from full-term (≥37 weeks) pregnancies. These seasonal variations were associated with the core circadian clock system, which may have implications for human health and provide further evidence for the placenta as a peripheral oscillator. Results also suggested seasonal changes in placental inflammation and hypoxia. The circadian system regulates many biological processes, and seasonal effects on the clock may alter placental function and fetal development. Further study of seasonal and circadian effects on the placenta could shed light on the pathways that influence the associations between season of birth and human health, as well as possible therapeutic targets.

## Data Availability

Archived placental transcriptomic data is available at dbGaP (phs001586.v1.p1)

## List of non-standard abbreviations

BH: Benjamini and Hochberg
DE: Differential expression
FDR: False Discovery Rate
FE: fall equinox
SE: spring equinox
SS: summer solstice
WS: winter solstice

## ACKNOWLEDGMENTS

This work was supported by funding from the National Institutes of Health (NIH-NIEHS R01ES022223 [to CJM, JC, KH], NIH-NIEHS R24ES025145 [to CJM], NIH-NIEHS P01 ES022832 [to CJM], NIH-NICHD F31 HD097918 [to DACT], and NIH-NIEHS T32 ES012870 [to DACT]) and by the United States Environmental Protection Agency (US EPA grant RD83544201 [to CJM]). The study sponsors did not have any role in the study design, collection, analysis, and interpretation of the data, the writing of the report, or the decision to submit the paper for publication.

## AUTHOR CONTRIBUTIONS

D. Clarkson-Townsend designed the research, performed the research, analyzed the data, and wrote the manuscript; L. Kennedy contributed to data analysis and edited the manuscript; T. Everson contributed to data analysis and edited the manuscript; M. Deyssenroth curated the data and edited the manuscript; A. Burt curated the data and edited the manuscript; K. Hao curated the data and edited the manuscript; J. Chen curated the data and edited the manuscript; M. Pardue contributed to conceptualization and edited the manuscript; C. Marsit contributed to conceptualization and data analysis, curated the data, provided resources for data acquisition, and edited the manuscript.

---

**Supplementary Figure 1.**
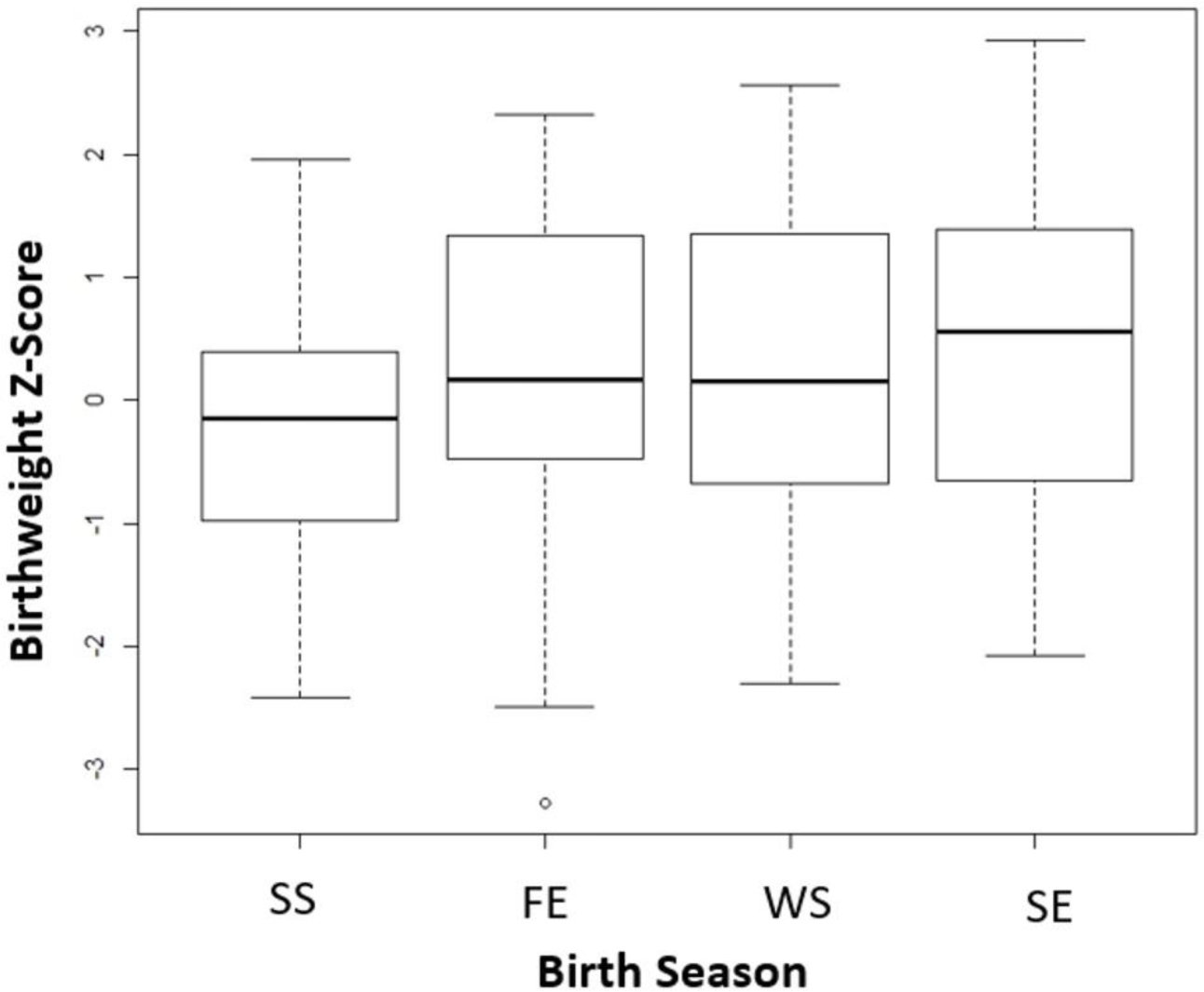
Boxplots of infant birthweight z-scores by season of birth.

**Supplementary Figure 2.**
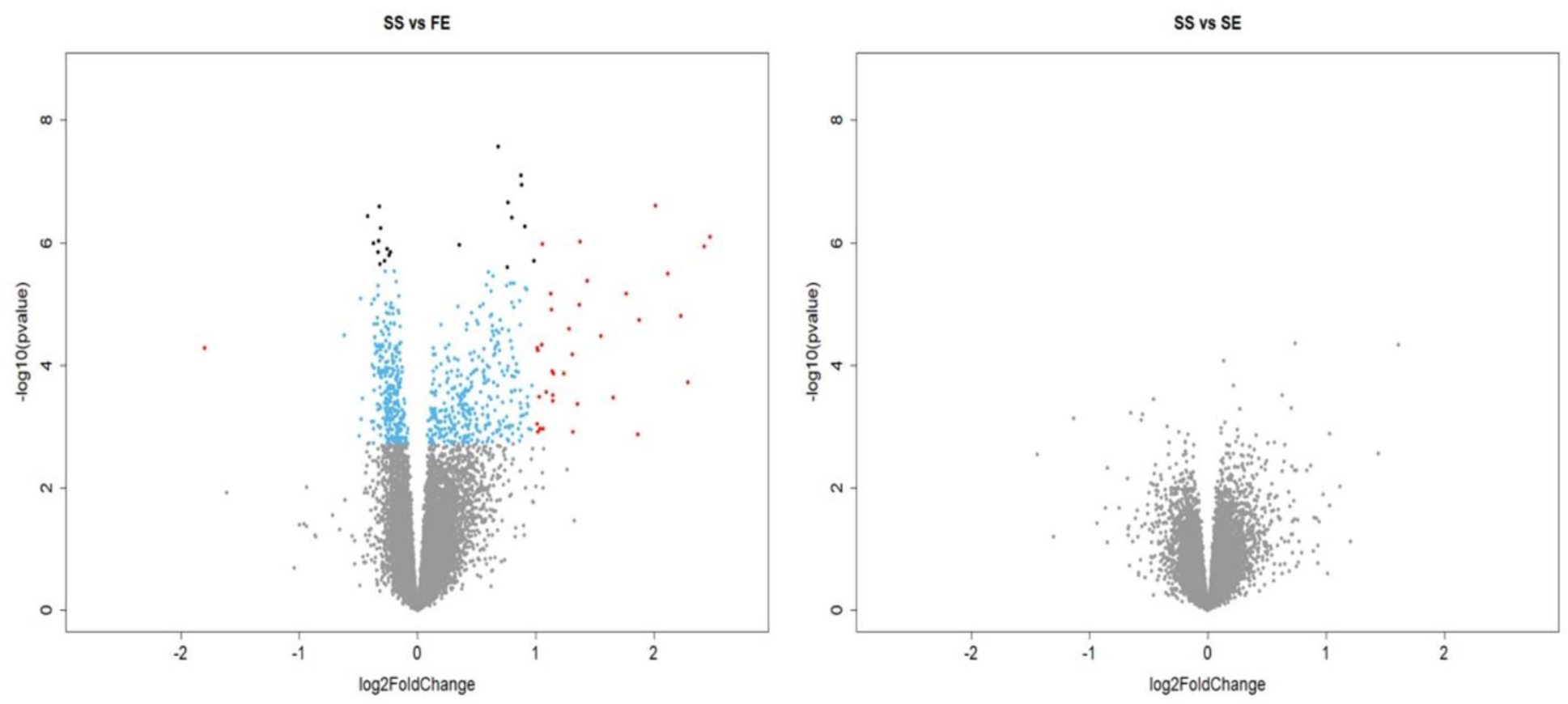
Volcano plot of results from the transcriptome wide association study of gene expression by birth season showing Wald test results comparing spring (SE) and fall (FE) to the summer (SS) referent group. Transcripts are plotted by effect size estimate [Log2(Fold Change), x-axis] and statistical significance [-Log10(Pvalue), y-axis], with grey dots indicating non-significant FDR q≥0.05, blue dots indicating FDR q<0.05, black dots indicating Bonferroni p<0.05, and red dots indicating absolute Log2(Fold Change)>1 and FDR q<0.05.

**Supplementary Figure 3.**
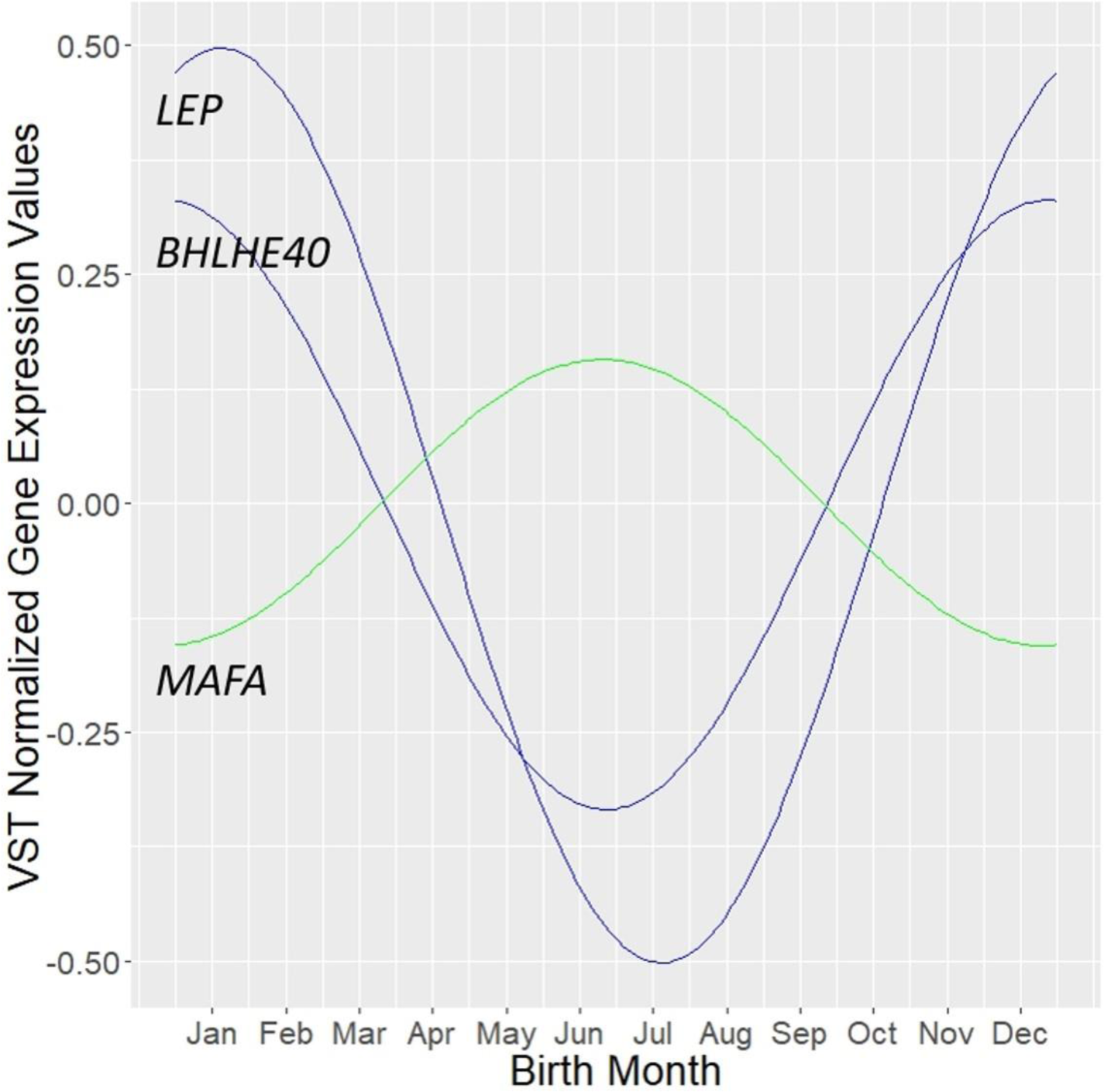
Graph showing the fitted curves BHLHE40, LEP, and MAFA when analyzed in the cosinor analysis.

**Supplementary Table 1.** LRT model results from DE analysis.

**Supplementary Table 2.** Wald test results from DE analysis.

**Supplementary Table 3.** Cosinor analysis results.

**Supplementary Table 4.** EnrichR results.

**Supplementary Table 5.** Transcripts (FDR q<0.05) that overlapped between the differential expression and cosinor analyses.

